# Effects of a single-dose methylphenidate challenge on resting-state functional connectivity in stimulant-treatment naive children and adults with ADHD

**DOI:** 10.1101/2022.02.04.22270336

**Authors:** Antonia Kaiser, Caroline Broeder, Jessica Cohen, Linda Douw, Liesbeth Reneman, Anouk Schrantee

## Abstract

Prior studies suggest that methylphenidate, the primary pharmacological treatment for attention-deficit/hyperactivity disorder (ADHD), alters functional brain connectivity. As the neurotransmitter systems targeted by methylphenidate undergo significant alterations throughout development, the effects of methylphenidate on functional connectivity may also be modulated by age. Therefore, we assessed the effects of a single methylphenidate challenge on brain network connectivity in stimulant-treatment naïve children and adults with ADHD. We obtained resting-state functional MRI from 50 boys (10-12 years of age) and 49 men (23-40 years of age) with ADHD (DSM IV, all subtypes), before and after an oral challenge with 0.5 mg/kg methylphenidate; and from 11 boys and 12 men as typically-developing controls. Connectivity strength (CS), eigenvector centrality (EC), and betweenness centrality (BC) were calculated for the striatum, thalamus, dorsal anterior cingulate cortex (dACC), and prefrontal cortex (PFC). In line with our hypotheses, we found that methylphenidate decreased measures of connectivity and centrality in the striatum and thalamus in children with ADHD, but increased the same metrics in adults with ADHD. Surprisingly, we found no major effects of methylphenidate in the dACC and PFC in either children or adults. Interestingly, pre-methylphenidate, participants with ADHD showed aberrant connectivity and centrality compared to controls predominantly in frontal regions. Our findings demonstrate that methylphenidate’s effects on connectivity of subcortical regions are age-dependent in stimulant-treatment naïve ADHD patients, likely due to ongoing maturation of dopamine and noradrenaline systems. These findings highlight the importance for future studies to take a developmental perspective when studying the effects of methylphenidate treatment.

## Introduction

In recent years, attention-deficit/hyperactivity disorder (ADHD) has been increasingly considered a disorder of brain-wide network dysconnectivity rather than of region-specific deficits [1,2]. Methylphenidate, the primary pharmacological treatment for ADHD, has been proposed to alter functional connectivity in various brain-wide functional circuits affected by ADHD [3]. For instance, normalized connectivity in fronto-parietal-cerebellar circuits has been observed in children with ADHD following acute methylphenidate. This was first observed by An et al., demonstrating that a single dose of methylphenidate compared to placebo, upregulated abnormally decreased local connectivity in bilateral ventral prefrontal cortices and the cerebellar vermis, and downregulated abnormally increased local connectivity in the right parietal and visual areas in children with ADHD [4]. Similarly, Silk et al. found that a single dose of methylphenidate compared to placebo normalized increased functional connectivity in occipital, temporal and cerebellar regions and visual, executive, and default mode networks in adolescents with ADHD [5]. More recently, alterations in fronto-parietal-cerebellar circuits have also been observed following prolonged methylphenidate treatment in medication-naïve children with ADHD [6]. Finally, preliminary evidence suggests that such a normalization might also occur in adults with ADHD [7,8]. However, due to methodological heterogeneity in previous studies, including prior use of stimulant medications, results remain inconclusive [3].

Methylphenidate acts by inhibiting dopamine and noradrenaline reuptake in the brain [9]. As the dopamine system undergoes significant alterations throughout development [10], methylphenidate-induced effects on functional connectivity may be modulated by age. For example, a recent longitudinal study demonstrated an age-dependent effect of prolonged stimulant treatment-response on cingulo-opercular network connectivity [11]. Moreover, exposure to stimulants during sensitive stages of maturation might cause developmental alterations, a process called neuronal imprinting [12]. Indeed, animal studies suggest that the age at initiation of methylphenidate-treatment affects its influence on development in a highly specific manner [13]. In the same sample as described here, we also observed that the effects of methylphenidate may be modulated by age; we found that acute methylphenidate decreased thalamic cerebral blood flow only in children, but not in adults [14]. Moreover, we observed that prolonged methylphenidate-treatment followed by an acute challenge with methylphenidate significantly influenced cerebral blood flow in the striatum and thalamus in children, but not adults, nor in the placebo conditions [15].

Here, we aimed to assess the effects of a single-dose challenge of methylphenidate on resting-state functional MRI (rs-fMRI) network connectivity in stimulant-treatment naïve children and adults with ADHD using graph theoretical measures, to investigate potential age-dependent neural mechanisms involved in stimulant-induced changes in ADHD. Based on previous findings, we expected significant methylphenidate-induced alterations in connectivity of the striatum, thalamus, dorsal anterior cingulate cortex (dACC), and prefrontal cortex (PFC). We expected that these effects would differ between children and adults because of functional maturation of the dopamine and noradrenaline system [10]. In addition, based on studies reporting altered connectivity in individuals with ADHD compared to typically-developing control participants, we hypothesized that an acute dose of methylphenidate would strengthen connectivity for these four brain regions in adults, whereas in children, we expected increased connectivity in frontal regions (PFC and dACC) and decreased connectivity in the thalamus and striatum.

## Methods

We included 50 stimulant treatment-naive boys (10-12 years of age) and 49 stimulant-treatment naive men (23-40 years of age) that were part of the “effects of Psychotropic drugs On the Developing brain - methylphenidate” (ePOD-MPH) trial (NTR3103 and NL34509.000.10; [15,16]). They were recruited through clinical programs at the Child and Adolescent Psychiatry Center Triversum (Alkmaar), the Department of Child and Adolescent Psychiatry at the Bascule/AMC (Amsterdam), and the PsyQ Mental Health Facility (The Hague). All participants were diagnosed with ADHD (DSM-IV, all subtypes) by an experienced psychiatrist, using a structured interview, (Diagnostic Interview Schedule for Children (NIMH-DISC-IV): authorized Dutch translation [17]) and the Diagnostic Interview for ADHD (DIVA 2.0) for adults [18]. In addition, as a typically-developing comparison group, we included 11 boys (aged 10–12 years) and 12 men (aged 23–40 years) as non-ADHD control participants, who received pre methylphenidate scans only (Table 1).

**Table 1.**
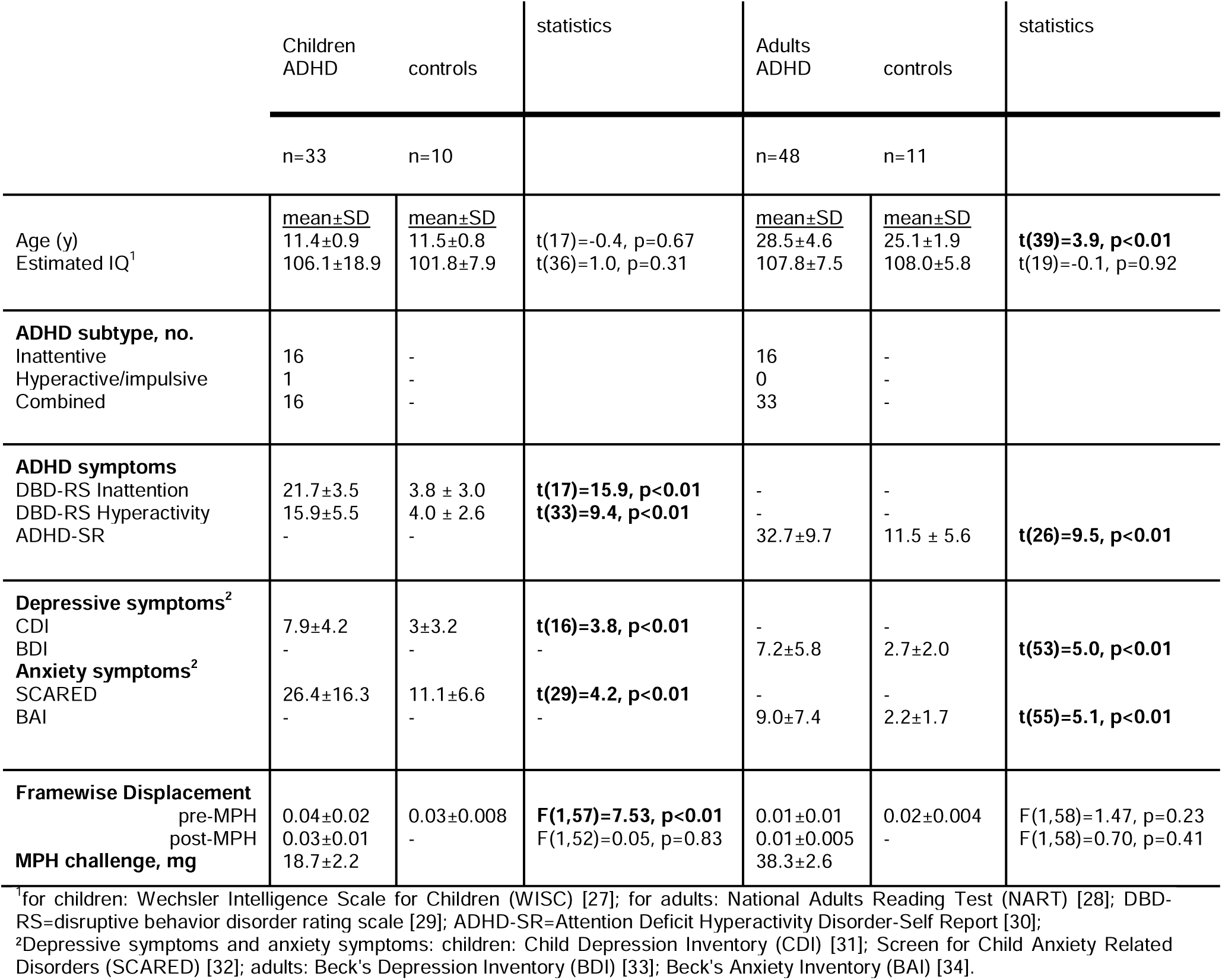
Characteristics of participants included in the rs-fMRI analysis. Significant effects are indicated in bold (p<0.05).

Exclusion criteria were: comorbid axis I psychiatric disorders requiring treatment with medication at study entry, a history of major neurological or medical illness or clinical treatment with drugs influencing the dopamine system (for adults before 23 years of age), such as stimulants, neuroleptics, antipsychotics, and/or D2/3 agonists (see Supplementary Material for more detail). The study was approved by the medical ethical committee and consequently monitored by the Clinical Research Unit of the Amsterdam University Medical Center, University of Amsterdam, Amsterdam, the Netherlands. All participants and parents or legal representatives of the children provided written informed consent.

The primary outcome measure of ePOD-MPH trial was to report on the modification by age of methylphenidate treatment on the outgrowth of the dopamine system by using pharmacologic MRI [15]. Here we report on acute effects of methylphenidate on the baseline rs-fMRI measurement of the trial, during which ADHD participants underwent two MRI scans, one before and one 90 min after an oral challenge of short-acting methylphenidate (Sandoz B.V., Weesp, the Netherlands; 0.5 mg/kg with a maximum of 20 mg in children and 40 mg in adults). The dose was chosen so that 80% of dopamine transporters were occupied [19], and we chose 90 min of waiting period for optimal occupation of these transporters. Typically developing control subjects did not receive a challenge of methylphenidate.

## Resting-state fMRI

Data were acquired on 3T Philips scanners (Philips Healthcare, Best, The Netherlands) using an 8-channel receive-only head coil. A 3D T1-weighted anatomical scan was acquired for registration purposes, and resting-state fMRI (rs-fMRI) data were acquired using a single-shot echo-planar imaging sequence (TR/TE=2300/30ms, resolution=2.3×2.3×3 mm, 39 sequential slices, FA=80°, dynamics=130).

Preprocessing was performed using FMRIPREP v1.2.3 [18; RRID: SCR_016216], including ICA-AROMA. Subsequently, white matter (WM) and cerebral spinal fluid (CSF) signals (obtained before ICA-AROMA) were regressed out and high-pass-filtering (100s) was applied using FSL. The Brainnetome atlas was used to define 246 parcels ([21]; Figure 1A+B) and time-series per participant were extracted and z-scored. Framewise displacement (FD) values were low-pass filtered to remove respiration artifacts [22] and timepoints where FD>0.2mm were scrubbed. Participants were excluded from further analyses if mean FD>0.2mm or if number of volumes after scrubbing ≤ 104.

Cleaned time series were then used to calculate connectivity matrices using Pearson correlations, resulting in a 246×246 connectivity matrix per participant, which was absolutized for further analyses (Figure 1C+D). Temporal signal-to-noise (tSNR) maps were calculated per participant to remove low-tSNR nodes.

**Figure 1.**
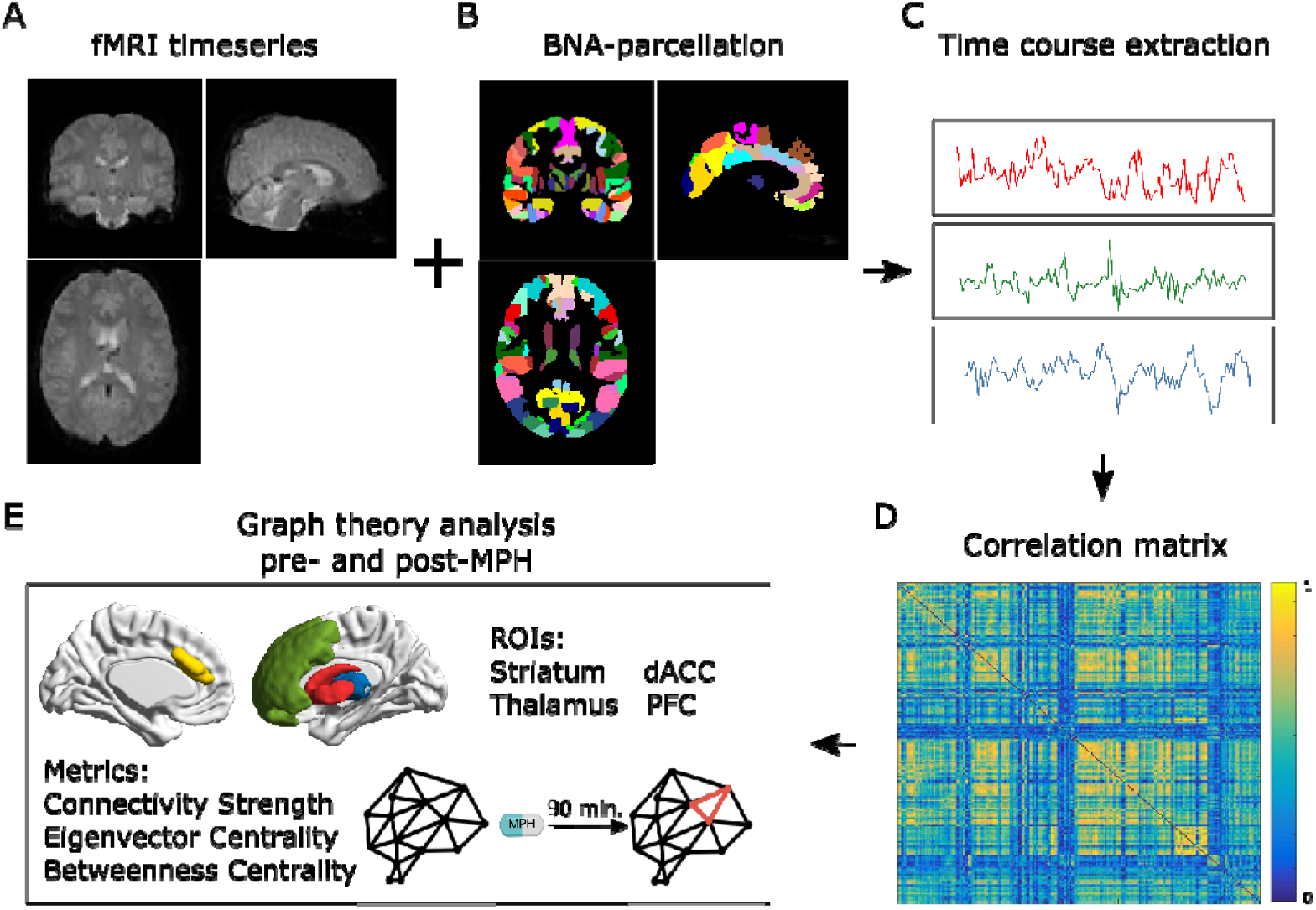
Analysis overview. **A+B)** To construct functional brain networks per participant, the Brainnetome atlas (BNA) was used to define 246 parcels [21]. **C+D)** The cleaned time series were then used to calculate connectivity matrices using Pearson correlations, resulting in a 246×246 connectivity matrix per participant, which was absolutized for further analyzes. **E)** Graph theory measures were calculated from the connectivity matrices using the Brain Connectivity Toolbox (Rubinov and Sporns 2010). Connectivity strength (CS), betweenness centrality (BC), and eigenvector centrality (EC) were calculated for four regions of interest (ROIs): striatum, thalamus, dorsal anterior cingulate cortex (dACC), and prefrontal cortex (PFC).

Graph theory measures were calculated for the whole brain from connectivity matrices using the Brain Connectivity Toolbox ([23]; [RRID:SCR_004841]; Figure 1E). Quality control measures as defined by Ciric et al. (2017), as well as the number of negative correlations and average correlation coefficients, were calculated (Supplementary Figure 1; Supplementary Table 1). Connectivity strength (CS), betweenness centrality (BC), and eigenvector centrality (EC) were calculated and consequently averaged for four regions of interest (ROIs): striatum, thalamus, dorsal anterior cingulate cortex (dACC), and prefrontal cortex (PFC) (Brainnetome region numbers per ROI in Supplementary Table 2). The striatum was selected because it is rich in dopamine transporters and is the primary target of methylphenidate. The thalamus and dACC were selected because animal literature has demonstrated the largest age-dependent effects of methylphenidate in these two important projections from the striatum [12]. Finally, the PFC was selected due to its hypothesized importance and its interconnection with other areas that are affected by ADHD [24]. Correlations of all connectivity measures and FD can be found in Supplementary Table 3. Further details on the analysis methods can be found in the Supplementary Material.

## Statistical Analysis

Statistical analyses were conducted using R v.3.5.3 [25]. All data were checked for normality and, in case of non-normality, log-transformed. Linear mixed-effects models were used to analyze changes in fMRI connectivity per age group separately to investigate the main effect of methylphenidate (pre- and post-challenge of acute methylphenidate) using the lme4 package [26]. Linear models were used to analyze the differences between the ADHD participants and controls at pre-methylphenidate. The average whole-brain CS per participant was added to the model as a covariate. FD and a variable representing the scanner that was used were tested as possible covariates, but not significant and thus not included in the models. Multiple comparison correction within modalities was performed using Sidak’ correction: α*=1-(1-α)^1/m^, with α=0.05 and m=4 (number of ROIs), which resulted in an α*=0.0127.

## Results

### Participants

Of the 99 ADHD patients scanned, data from 81 participants with ADHD and 21 typically-developing controls were analyzed (Table 1). One adult ADHD participant was excluded because of undisclosed prior stimulant treatment (more details about the trial are published elsewhere [15]). Seventeen children with ADHD were excluded due to excessive motion (whose characteristics did not differ from the included children (Supplementary Results)).

### Rs-fMRI connectivity

All results of the statistical tests, as well as the estimated means and 95% confidence intervals, can be found in Table 2.

**Table 2.**
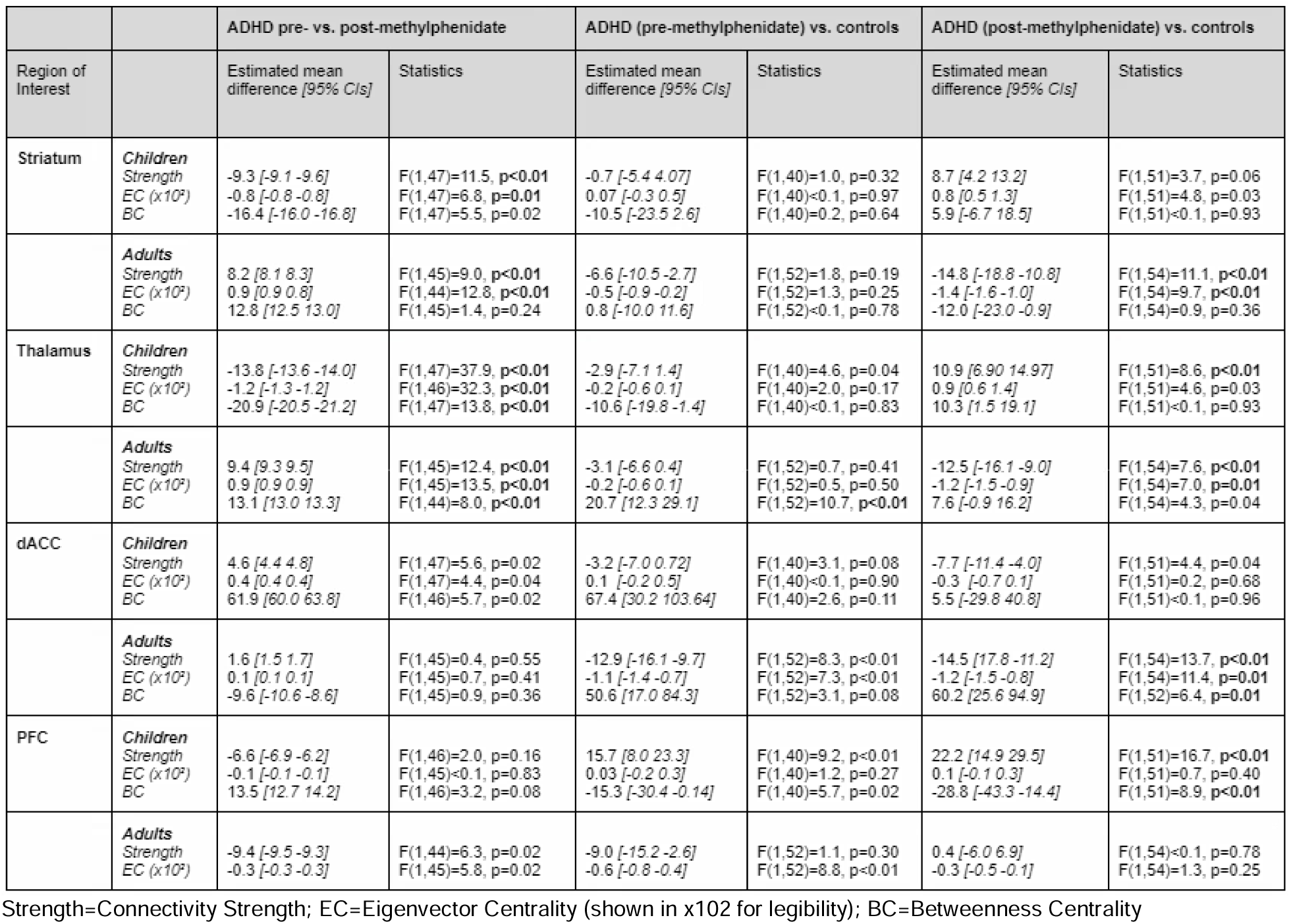
Results of statistical tests. Linear mixed-effects models were used to analyze changes in fMRI connectivity per age group separately to investigate the main effect of methylphenidate (pre- and post-challenge of acute methylphenidate). Linear models were used to analyze the differences between the ADHD participants and controls at pre- and post-methylphenidate. The average whole-brain CS per participant was added to the model as a covariate. Multiple comparison correction within modalities was performed using Sidak’s correction, which resulted in an α*=0.0127. Significant effects are indicated in bold.

#### Striatum

Pre- to post-methylphenidate, in children with ADHD, CS, and EC significantly decreased, but changes in BC did not survive multiple comparison corrections. Pre- to post-methylphenidate, in adults with ADHD, the opposite effect was found; both CS and EC significantly increased, but BC did not change significantly.

Pre-methylphenidate, neither children nor adults with ADHD differed significantly from the respective controls in any of the connectivity metrics. Post-methylphenidate, in children with ADHD, none of the connectivity metrics differed significantly from the respective young controls. Post-methylphenidate, in adults with ADHD, CS and EC differed significantly from the adult controls, but BC did not differ significantly. (Figure 2A; Table 2).

**Figure 2.**
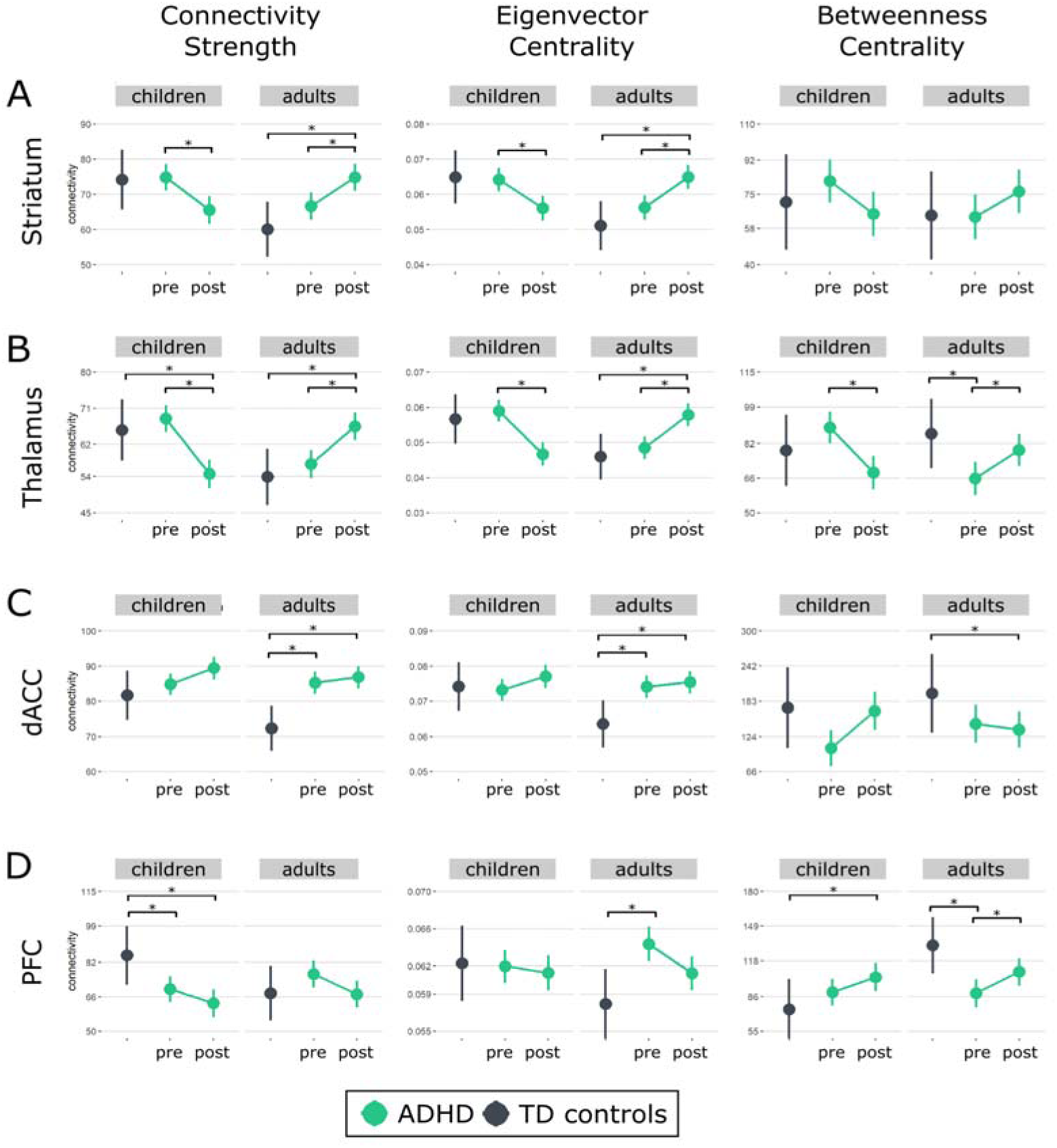
Functional connectivity within the ROIs. Connectivity strength, eigenvector centrality, and betweenness centrality are shown for the **A)** striatum **B)** thalamus **C)** dorsal anterior cingulate cortex (dACC) and **D)** prefrontal cortex (PFC). Estimated marginal means and 95% confidence intervals at pre-MPH (pre) and post-MPH (post) for children and adults are shown in green. Estimated means and 95% confidence intervals for TD controls are shown in black. Significant effects are indicated with an asterisk (*; p<0.0127).

#### Thalamus

Pre- to post-methylphenidate, in children with ADHD, CS, EC and BC decreased significantly. Pre- to post-methylphenidate, in adults with ADHD, the opposite effect was found; CS, EC and BC increase significantly. Pre-methylphenidate, children with ADHD did not differ significantly from the young controls in any of the connectivity metrics.

Pre-methylphenidate, adult ADHD participants showed lower BC than adult controls, but CS and EC did not differ significantly. Post-methylphenidate, in children with ADHD, CS was significantly different from the respective controls, but EC and BC were not significantly different. Post-methylphenidate, in adults with ADHD, CS and EC were significantly different from the respective controls (Figure 2B; Table 2).

#### dACC

Pre- to post-methylphenidate, in children with ADHD, CS, BC and EC changes did not survive multiple comparison corrections. Pre- to post-methylphenidate, in adults with ADHD, none of the connectivity metrics changed significantly.

Pre-methylphenidate, children with ADHD did not differ significantly from the respective controls in any of the connectivity metrics. Pre-methylphenidate, adult ADHD participants showed higher CS and EC values than the adult controls, but BC did not differ significantly. Post-methylphenidate, in children with ADHD, none of the connectivity metrics differed from the controls. Post-methylphenidate, in adults with ADHD, CS, EC and BC differed significantly from (Figure 2C;Table 2).

#### PFC

Pre- to post-methylphenidate, in children with ADHD, none of the graph-theory metrics changed significantly. Pre- to post-methylphenidate, in adults with ADHD, CS and EC did not survive multiple comparison corrections, BC was found to increase significantly.

Pre-methylphenidate, children with ADHD showed significantly higher CS values than controls. None of the other connectivity measures differed significantly. Adults with ADHD showed no differences to controls in CS, but significantly higher EC values and lower BC values than the control group. Post-methylphenidate, in children with ADHD, EC was not significantly different from the young controls, but CS and BC were found to be significantly different. Post-methylphenidate, in adults with ADHD, none of the connectivity metrics were significantly different from the adult controls (Figure 2D; Table 2).

## Discussion

The goal of this study was to investigate the effects of acute methylphenidate on rs-fMRI connectivity in stimulant-treatment naïve children and adults with ADHD. In line with our hypotheses, we found that methylphenidate decreased measures of connectivity and centrality in subcortical ROIs in children with ADHD, but increased the same metrics in adults with ADHD, indicating an age-dependent acute effect of methylphenidate in dopamine-sensitive regions. Surprisingly, we found no major effects of methylphenidate in frontal ROIs in either children or adults. Interestingly, at pre-methylphenidate, participants with ADHD showed aberrant connectivity and centrality predominantly in frontal ROI compared to controls.

### Effect of methylphenidate in children with ADHD

A recent review on the effects of stimulant medication on rs-fMRI connectivity in individuals with ADHD shows that methylphenidate appears to modulate several rs-fMRI networks, but the number of studies is small, and the results are heterogeneous [3]. In line with findings from Silk et al. [5], we observed that acute methylphenidate decreased connectivity in the striatum and thalamus, whereas in the dACC we found non-significant increases in connectivity after a single dose of methylphenidate. This is in agreement with a previous study reporting that acute methylphenidate increased connectivity in frontal regions [4]. Notwithstanding, our study has some methodological differences compared to previous studies. Firstly, all our participants were stimulant-treatment naïve, whereas in other studies medication status was inconsistent. Therefore, our study rules out the influence of prior medication on connectivity through prolonged effects of stimulants on the dopamine system. For example, prolonged MPH treatment has been shown to impact (proxy measures of) dopamine function in juvenile animals and children [12,15,35]. Furthermore, long-term stimulant treatment normalized delayed structural maturation of the PFC in individuals with ADHD, which may reflect dopaminergic adaptive processes [36,37].

Secondly, we assessed graph theory metrics, whereas Silk et al. [5] using Network Based Statistics to identify connections that are affected by methylphenidate, and An et al. [4] used regional homogeneity, reflecting local synchronized brain activity, considered to be a measure of functional segregation [38]. As such, our study extends prior literature from connectivity metrics to topology metrics, which allows us to not only assess individual nodes or global connectivity, but to assess the importance and integration of pre-specified nodes *within* the global network. In subcortical regions, methylphenidate affects average connectivity (CS) and nodal importance (EC), suggesting changes in the role of these regions in both local and global network topology. In frontal regions on the other hand, we observe marginal increases in global importance (BC) following methylphenidate, which might indicate a more important role for these regions regarding information flow in the network [39,40]. Thirdly, both previous studies included placebo conditions, whereas we used a pre-post design. Finally, the dose that we used was slightly higher than Silk et al. (2017; 0.41mg/kg) and substantially higher than An et al. (10 mg), which may have affected functional connectivity differently [4], particularly considering the inverted-U relationship between dopamine levels and cognition [41,42].

Although previous studies have reported that methylphenidate normalizes brain activity [43,44] and connectivity [4], our results do not support these findings. Instead, pre-methylphenidate, we show no group differences in connectivity in subcortical ROIs, and our findings suggest that methylphenidate-induced changes in connectivity deviate from the control-like state. We could speculate that these discrepancies are due to divergent brain development in ADHD, affecting local vs. global metrics differently. As such, methylphenidate could normalize local connectivity and activity, as demonstrated by previous studies, but compensate for altered network structure on a global level, as found here. In the PFC on the other hand, we found higher CS compared to controls, one of the latest brain regions to mature [45]. This is partly in line with two recent meta-analyses proposing increased connectivity within the executive control network in children with ADHD [46,47], potentially reflecting greater mental effort to compensate for executive function in ADHD.

### Effect of methylphenidate in adults with ADHD

The present study is, to our knowledge, the first to investigate the acute effects of methylphenidate in stimulant-treatment naive adults with ADHD. In agreement with our hypotheses, our findings indicate that methylphenidate increased overall connectivity and importance of striatal and thalamic nodes within the brain network. Our results show overlap with regions identified in a study investigating prolonged effects of methylphenidate in adults [7], and correspond to findings from typically-developing adults showing that acute methylphenidate increased connectivity between the thalamus and attention networks, and subcortical regions [48,49]. These findings, together with the absence of major differences in connectivity when compared to controls, suggest that the mechanisms underlying the effects of methylphenidate on subcortical connectivity are largely comparable between adults with and without ADHD. However, this is in contrast with evidence from Positron Emission Tomography (PET) studies reporting significant differences in striatal dopamine release between adults with ADHD and controls following a stimulant challenge; albeit in different directions [50,51]. Together, this suggests that differential effects of methylphenidate on subcortical dopamine release may not directly translate into differential subcortical connectivity between individuals with ADHD and controls.

The pattern observed in cortical regions is more complex. In the dACC, methylphenidate did not induce changes in connectivity in participants with ADHD, despite higher pre-methylphenidate connectivity compared to controls. Such hyperconnectivity [52] could be speculated to be a result of developed compensatory processes, in response to reduced network efficiency [53], particularly in adults who were never treated with ADHD medication. Interestingly, the absence of normalized dACC after methylphenidate could suggest that such processes are dopamine and noradrenaline-independent. Alternatively, individual differences may be too large to observe group differences, or such processes affect other network measures than those studied here. Conversely, in the PFC, we found that BC increased, whereas CS and EC decreased after methylphenidate. This would mean that methylphenidate increases the role of the PFC as a global communication hub (i.e., BC), but reduces connectivity of the PFC with other regions (i.e., CS and EC); meaning that the PFC connections become more specialized for network communication.

### Age-dependent effects of methylphenidate in ADHD

The effects of methylphenidate on the brain have been proposed to be age-dependent [11–13]. Indeed, we previously showed that thalamic cerebral blood flow was reduced following acute methylphenidate in children, but not adults with ADHD [15]. Accordingly, we here find an opposite effect of acute methylphenidate in thalamic and striatal connectivity in children compared to adults. Nevertheless, these age-effects may not be specific to ADHD, as functional connectivity changes over development, complicating intergenerational comparisons [54]. Functional segregation appears predominant in children, whereas functional integration prevails in adults. As such, typical development of functional connectivity is characterized by simultaneous reduction of local circuitry and strengthening of long-range connectivity [55,56]. Nevertheless, we can speculate that the difference in methylphenidate-induced connectivity changes between children and adults might result from maturation of dopaminergic and noradrenergic systems [10]. For instance, adults display a more segregated architecture in the fronto-parietal network, including the dorsal basal ganglia (i.e., caudate nucleus) [57], possibly through changes in the dopamine system in the frontal cortex [58–60]. This network is, e.g., important for the top-down regulation of emotion and attention [61]. Indeed, a recent longitudinal study on the effects of stimulant treatment response and age found a significant influence on cingulo-opercular network connectivity [11]. The age-dependent effects on striatal and thalamic connectivity reported here could therefore be due to compensatory mechanisms taking place in the adults, especially given that they were stimulant-treatment naïve before the study. It has been argued that the neuropathology of childhood remittent cases could be attributed largely to a delayed frontal cortex maturation, whereas the neuropathology of persistent cases is linked more to pathology in extra-frontal and subcortical structures [62]. In summary, this suggests that the efficacy of stimulant therapy may not be based on normalization only, but rather depend on combinations of factors that return the network organization to typical topology for some systems while reorganizing others. In other words, it might be that altered networks in the brain do not need to return to the control state to function in the desired way, a restructuring of function could be sufficient. It is therefore important that future studies take age-dependent effects into account.

In addition, previous studies have suggested potential neural differences between persistent and remitted adults with ADHD [63]. By definition, our adult ADHD sample had persistent ADHD, whereas this remains to be assessed for our pediatric sample [64,65]. Longitudinal (f)MRI studies on ADHD persisters and remitters with childhood ADHD will be crucial to gain more insight into the differences in brain connectivity of persisting and remitting ADHD in childhood [66]. Speculatively, in addition to developmental differences, our results may partially be explained by neuronal differences between these two ADHD phenotypes. Norman et al., indeed found reduced connectivity within the inferior frontal gyrus in children with ADHD to be indicative of longitudinal risk for ADHD inattention symptoms [67]. Additionally, because we included stimulant-treatment naïve individuals with ADHD, the adults might not represent a typical sample, as most adults with ADHD will receive medication before adulthood. For a long time it has been debated if ADHD may also be developed in adulthood, with no previous symptoms in childhood (“adult-onset ADHD”; [68]). However, a recent review argues that symptoms in adults indeed exist but that their source would be either symptoms that were previously surpassed, were not properly assessed before, or not detected earlier [69].

One of the main strengths of this study is that we included both stimulant-treatment naïve boys and men with ADHD and that, compared to previous studies on the acute effects of methylphenidate, we included a larger number of participants. However, limitations of our study are that the results cannot be extrapolated to all children and adults with ADHD, because we only studied participants with restricted age ranges. Furthermore, we included only male participants to reduce heterogeneity, but this limits the generalizability to female participants. Additional studies are needed in females, since female sex hormones modulate dopamine transporter expression [70]. Furthermore, the comparisons between participants with ADHD and control participants have to be interpreted with caution, due to the small control groups and because control participants did not receive a methylphenidate challenge. Ethical considerations did not permit us to administer methylphenidate to the young controls, therefore, the controls were assessed only once.

## Conclusion

Taken together, in line with our hypothesis, we found opposing effects of acute methylphenidate on connectivity strength and the relative importance of the nodes in subcortical regions, in children compared to adults. In contrast to what we expected, MPH-induced changes in connectivity of frontal cortical regions were marginal. They did not indicate differences between age groups, and mainly global importance of these regions (i.e., their importance as a hub) within the network was increased. Therefore, we conclude that acute methylphenidate-effects on connectivity measures in dopamine-sensitive subcortical, but not cortical regions, are different in children and adults with ADHD, possibly due to changes of the dopamine and noradrenergic systems during maturation. These findings highlight the importance for future studies to investigate the age-dependent effects of long-term methylphenidate treatment, ideally in previously medication-naive individuals, on graph-theoretical connectivity measures, with a focus on centrality measures of subcortical regions. Additionally, we did not find normalizing effects of acute methylphenidate in either of the age groups, indicating that the previously found normalization towards a control state might be present on the local connectivity level, whereas on the global network level methylphenidate may give rise to reorganization of function.

## Data Availability

All data produced in the present study are available upon reasonable request to the authors

## Acknowledgments

We want to thank all participants and their parents for participating in this study and all students who helped collect and analyze the data.

## Author contributions statement

All authors made a substantial contribution to the concept and design, acquisition of data or analysis and interpretation of data, drafted the article or revised it critically for important intellectual content, and approved the version to be published.

## References

1. Castellanos FX, Proal E. Large-scale brain systems in ADHD: Beyond the prefrontal-striatal model. Trends Cogn Sci. 2012.

2. Samea F, Soluki S, Nejati V, Zarei M, Cortese S, Eickhoff SB, et al. Brain alterations in children/adolescents with ADHD revisited: A neuroimaging meta-analysis of 96 structural and functional studies. Neurosci Biobehav Rev. 2019;100:1–8.

3. Pereira-Sanchez V, Franco AR, Vieira D, de Castro-Manglano P, Soutullo C, Milham MP, et al. Systematic Review: Medication Effects on Brain Intrinsic Functional Connectivity in Patients With Attention-Deficit/Hyperactivity Disorder. J Am Acad Child Adolesc Psychiatry. 2020;60:222–235.

4. An L, Cao XH, Cao QJ, Sun L, Yang L, Zou QH, et al. Methylphenidate normalizes resting-state brain dysfunction in boys with attention deficit hyperactivity disorder. Neuropsychopharmacology. 2013;38:1287–1295.

5. Silk TJ, Malpas C, Vance A, Bellgrove MA. The effect of single-dose methylphenidate on resting-state network functional connectivity in ADHD. Brain Imaging Behav. 2017;11:1422–1431.

6. Yoo JH, Kim D, Choi J, Jeong B. Treatment effect of methylphenidate on intrinsic functional brain network in medication-naïve ADHD children: A multivariate analysis. Brain Imaging Behav. 2018;12:518–531.

7. Cary RP, Ray S, Grayson DS, Painter J, Carpenter S, Maron L, et al. Network structure among brain systems in adult ADHD is uniquely modified by stimulant administration. Cereb Cortex. 2017;27:3970–3979.

8. Picon FA, Sato JR, Anés M, Vedolin LM, Mazzola AA, Valentini BB, et al. Methylphenidate Alters Functional Connectivity of Default Mode Network in Drug-Naive Male Adults With ADHD. J Atten Disord. 2020;24:447–455.

9. Cortese S, D’Acunto G, Konofal E, Masi G, Vitiello B. New Formulations of Methylphenidate for the Treatment of Attention-Deficit/Hyperactivity Disorder: Pharmacokinetics, Efficacy, and Tolerability. CNS Drugs. 2017;31:149–160.

10. Chen YI, Choi JK, Xu H, Ren J, Andersen SL, Jenkins BG. Pharmacologic neuroimaging of the ontogeny of dopamine receptor function. Dev Neurosci. 2010;32:125–138.

11. Norman LJ, Sudre G, Bouyssi-Kobar M, Sharp W, Shaw P. A Longitudinal Study of Resting-State Connectivity and Response to Psychostimulant Treatment in ADHD. Am J Psychiatry. 2021;178:744–751.

12. Andersen SL. Stimulants and the developing brain. Trends Pharmacol Sci. 2005;26:237–243.

13. Canese R, Adriani W, Marco EM, De Pasquale F, Lorenzini P, De Luca N, et al. Peculiar response to methylphenidate in adolescent compared to adult rats: A phMRI study. Psychopharmacology (Berl). 2009;203:143–153.

14. Schrantee A, Mutsaerts HJMM, Bouziane C, Tamminga HGH, Bottelier MA, Reneman L. The age-dependent effects of a single-dose methylphenidate challenge on cerebral perfusion in patients with attention-deficit/hyperactivity disorder. NeuroImage Clin. 2017;13:123–129.

15. Schrantee A, Tamminga HGHH, Bouziane C, Bottelier MA, Bron EE, Mutsaerts H-JJMMM, et al. Age-Dependent Effects ofMethylphenidate on the Human Dopaminergic System in Young vs Adult Patients With Attention-Deficit/Hyperactivity Disorder. JAMA Psychiatry. 2016;73:955–962.

16. Bottelier MA, Schouw MLJ, Klomp A, Tamminga HGH, Schrantee AGM, Bouziane C, et al. The effects of Psychotropic drugs On Developing brain (ePOD) study: Methods and design. BMC Psychiatry. 2014;14:48.

17. Ferdinand R, van der Ende J. DISC-IV Diagnostic Interview Schedule for Children [Dutch translation NIMH-DISC-IV]. Rotterdam, the Netherlands: Afdeling Kinderen Jeugdpsychiatrie, Sophia Kinderziekenhuis/Academisch Ziekenhuis Rotterdam; 1998.

18. Kooij SJJ, Boonstra AM, Swinkels SHN, Bekker EM, de Noord I, Buitelaar JK, et al. Reliability, validity, and utility of instruments for self-report and informant report concerning symptoms of ADHD in adult patients. J Atten Disord. 2008;11:445–458.

19. Swanson JM, Volkow ND. Serum and brain concentrations of methylphenidate: implications for use and abuse. Neurosci Biobehav Rev. 2003;27:615–621.

20. Esteban O, Ciric R, Finc K, Blair RW, Markiewicz CJ, Moodie CA, et al. Analysis of task-based functional MRI data preprocessed with fMRIPrep. Nat Protoc. 2020:1–17.

21. Fan L, Li H, Zhuo J, Zhang Y, Wang J, Chen L, et al. The Human Brainnetome Atlas: A New Brain Atlas Based on Connectional Architecture. Cereb Cortex. 2016;26:3508–3526.

22. Fair DA, Miranda-Dominguez O, Snyder AZ, Perrone A, Earl EA, Van AN, et al. Correction of respiratory artifacts in MRI head motion estimates. Neuroimage. 2020;208:116400.

23. Rubinov M, Sporns O. Complex network measures of brain connectivity: uses and interpretations. Neuroimage. 2010;52:1059–1069.

24. Mehta TR, Monegro A, Nene Y, Fayyaz M, Bollu PC. Neurobiology of ADHD: A Review. Curr Dev Disord Reports. 2019;6:235–240.

25. R Development Core Team RFFSC. R: A language and environment for statistical computing. 2011. 2011.

26. Bates D, Mächler M, Bolker B, Walker S. Fitting Linear Mixed-Effects Models Using lme4. J Stat Softw. 2015;67:1–48.

27. Kort W, Compaan EL, Bleichrodt N, Resing WCM, Schittekatte M, Bosmans M. WISC-III NL. London Psychol Corp. 2002. 2002.

28. Schmand B, Lindeboom J, van Harskamp F. Dutch Adult Reading Test. Lisse Swets En Zeitlinger. 1992. 1992.

29. Pelham Jr. WE, Gnagy EM, Greenslade KE, Milich R. Teacher ratings of DSM-III-R symptoms for the disruptive behavior disorders. J Am Acad Child Adolesc Psychiatry. 1992;31:210–218.

30. Kooij J. Adult ADHD: Diagnostic assessment and treatment. London: Springer-Verlag; 2012.

31. Kovacs M. The Children’s Depression Inventory (CDI). Psychopharmacol Bull. 1985;21:995–998.

32. Muris P, Merckelbach H, Van Brakel A, Mayer B, Van Dongen L. The screen for child anxiety related emotional disorders (SCARED): Relationship with anxiety and depression in normal children. Pers Individ Dif. 1998;24:451–456.

33. Beck AT, Ward CH, Mendelson M, Mock J, Erbaugh J. An inventory for measuring depression. Arch Gen Psychiatry. 1961;4:561–571.

34. Beck AT, Epstein N, Brown G, Steer RA. An inventory for measuring clinical anxiety: psychometric properties. J Consult Clin Psychol. 1988;56:893–897.

35. Moll GH, Hause S, Rüther E, Rothenberger A, Huether G, Al MET. Early Methylphenidate Administration to Young Rats. J Child Adolesc Psychopharmacol. 2001;11:15–24.

36. Shaw P, Sharp WS, Morrison M, Eckstrand K, Greenstein DK, Clasen LS, et al. Psychostimulant treatment and the developing cortex in attention deficit hyperactivity disorder. Am J Psychiatry. 2009;166:58–63.

37. Castellanos XF, Patti LP, Sharp W, Jeffries NO, Greenstein DK, Clasen LS, et al. Developmental trajectories of brain volume abnormalities in children and adolescents with attention-deficit/hyperactivity disorder. J Am Med Assoc. 2002;288:1740–1748.

38. Lv H, Wang Z, Tong E, Williams LM, Zaharchuk G, Zeineh M, et al. Resting-state functional MRI: Everything that nonexperts have always wanted to know. Am J Neuroradiol. 2018;39:1390–1399.

39. Wang J, Zuo X, He Y. Graph-based network analysis of resting-state functional MRI. Front Syst Neurosci. 2010;4.

40. Farahani F V., Karwowski W, Lighthall NR. Application of Graph Theory for Identifying Connectivity Patterns in Human Brain Networks: A Systematic Review. Front Neurosci. 2019;0:585.

41. Arnsten AFT, Rubia K. Neurobiological circuits regulating attention, cognitive control, motivation, and emotion: Disruptions in neurodevelopmental psychiatric disorders. J Am Acad Child Adolesc Psychiatry. 2012.

42. Froudist-Walsh S, Bliss DP, Ding X, Jankovic-Rapan L, Niu M, Knoblauch K, et al. A dopamine gradient controls access to distributed working memory in monkey cortex. BioRxiv. 2020:2020.09.07.286500.

43. Czerniak SM, Sikoglu EM, King JA, Kennedy DN, Mick E, Frazier J, et al. Areas of the Brain Modulated by Single-Dose Methylphenidate Treatment in Youth with ADHD During Task-Based fMRI: A Systematic Review. Harv Rev Psychiatry. 2013;21:151.

44. Rubia K. ‘Cool’ inferior frontostriatal dysfunction in attention-deficit/hyperactivity disorder versus ‘hot’ ventromedial orbitofrontal-limbic dysfunction in conduct disorder: A review. Biol Psychiatry. 2011.

45. Mills KL, Goddings AL, Clasen LS, Giedd JN, Blakemore SJ. The developmental mismatch in structural brain maturation during adolescence. Dev Neurosci. 2014;36:147–160.

46. Gao Y, Shuai D, Bu X, Hu X, Tang S, Zhang L, et al. Impairments of large-scale functional networks in attention-deficit/hyperactivity disorder: A meta-analysis of resting-state functional connectivity. Psychol Med. 2019;49:2475–2485.

47. Sutcubasi B, Metin B, Kurban MK, Metin ZE, Beser B, Sonuga-Barke E. Resting-state network dysconnectivity in ADHD: A system-neuroscience-based meta-analysis. World J Biol Psychiatry. 2020;0:1–11.

48. Mueller S, Costa A, Keeser D, Pogarell O, Berman A, Coates U, et al. The effects of methylphenidate on whole brain intrinsic functional connectivity. Hum Brain Mapp. 2014;35:5379–5388.

49. Farr OM, Zhang S, Hu S, Matuskey D, Abdelghany O, Robert T, et al. The effects of methylphenidate on resting-state striatal., thalamic and global functional connectivity in healthy adults. Int J Neuropsychopharmacol. 2014;17:1177–1191.

50. Cherkasova M V, Faridi N, Casey KF, O’Driscoll GA, Hechtman L, Joober R, et al. Amphetamine-Induced Dopamine Release and Neurocognitive Function in Treatment-Naive Adults with ADHD. Neuropsychopharmacol 2014 396. 2013;39:1498–1507.

51. Volkow ND, Wang G-J, Newcorn J, Telang F, Solanto M V, Fowler JS, et al. Depressed dopamine activity in caudate and preliminary evidence of limbic involvement in adults with attention-deficit/hyperactivity disorder. Arch Gen Psychiatry. 2007;64:932–940.

52. Guo X, Yao D, Cao Q, Liu L, Zhao Q, Li H, et al. Shared and distinct resting functional connectivity in children and adults with attention-deficit/hyperactivity disorder. Transl Psychiatry 2020 101. 2020;10:1–12.

53. Konrad K, Eickhoff SB. Is the ADHD brain wired differently? A review on structural and functional connectivity in attention deficit hyperactivity disorder. Hum Brain Mapp. 2010;31:904–916.

54. Tooley UA, Bassett DS, Mackey AP. Environmental influences on the pace of brain development. Nat Rev Neurosci 2021 226. 2021;22:372–384.

55. Supekar K, Uddin LQ, Prater K, Amin H, Greicius MD, Menon V. Development of functional and structural connectivity within the default mode network in young children. Neuroimage. 2010;52:290–301.

56. Grayson DS, Fair DA. Development of large-scale functional networks from birth to adulthood: A guide to the neuroimaging literature. Neuroimage. 2017;160:15–31.

57. Fair DA, Cohen AL, Power JD, Dosenbach NUF, Church JA, Miezin FM, et al. Functional Brain Networks Develop from a “Local to Distributed” Organization. PLOS Comput Biol. 2009;5:e1000381.

58. Lidow M, Goldman-Rakic P, Gallager D, Rakic P. Distribution of dopaminergic receptors in the primate cerebral cortex: quantitative autoradiographic analysis using [3H]raclopride, [3H]spiperone and [3H]SCH23390. Neuroscience. 1991;40:657–671.

59. Lidow M, Rakic P. Scheduling of monoaminergic neurotransmitter receptor expression in the primate neocortex during postnatal development. Cereb Cortex. 1992;2:401–416.

60. Rosenberg D, Lewis, DA. Changes in the dopaminergic innervation of monkey prefrontal cortex during late postnatal development: a tyrosine hydroxylase immunohistochemical study. Biol Psychiatry. 1994;36:272–277.

61. Zhou Y, Liang M, Jiang T, Tian L, Liu Y, Liu Z, et al. Functional dysconnectivity of the dorsolateral prefrontal cortex in first-episode schizophrenia using resting-state fMRI. Neurosci Lett. 2007;417:297–302.

62. Francx W, Oldehinkel M, Oosterlaan J, Heslenfeld D, Hartman CA, Hoekstra PJ, et al. The executive control network and symptomatic improvement in attention-deficit/hyperactivity disorder. Cortex. 2015;73:62–72.

63. Mattfeld AT, Gabrieli JDE, Biederman J, Spencer T, Brown A, Kotte A, et al. Brain differences between persistent and remitted attention deficit hyperactivity disorder. Brain. 2014;137:2423–

64. Kessler RC, Adler LA, Barkley R, Biederman J, Conners CK, Faraone S V., et al. Patterns and Predictors of Attention-Deficit/Hyperactivity Disorder Persistence into Adulthood: Results from the National Comorbidity Survey Replication. Biol Psychiatry. 2005;57:1442–1451.

65. Caye A, Spadini A V., Karam RG, Grevet EH, Rovaris DL, Bau CHD, et al. Predictors of persistence of ADHD into adulthood: a systematic review of the literature and meta-analysis. Eur Child Adolesc Psychiatry 2016 2511. 2016;25:1151–1159.

66. Rubia K. Cognitive Neuroscience of Attention Deficit Hyperactivity Disorder (ADHD) and Its Clinical Translation. Front Hum Neurosci. 2018;12:1–23.

67. Norman LJ, Sudre G, Bouyssi-Kobar M, Sharp W, Shaw P. An examination of the relationships between attention/deficit hyperactivity disorder symptoms and functional connectivity over time. Neuropsychopharmacology. 2021;0:1–7.

68. Castellanos FX. Is Adult-Onset ADHD a Distinct Entity? Am J Psychiatry. 2015;172:929–931.

69. Taylor LE, Kaplan-Kahn EA, Lighthall RA, Antshel KM. Adult-Onset ADHD: A Critical Analysis and Alternative Explanations. Child Psychiatry Hum Dev 2021. 2021:1–19.

70. Wagner AK, Kline AE, Ren D, Willard LA, Wenger MK, Zafonte RD, et al. Gender associations with chronic methylphenidate treatment and behavioral performance following experimental traumatic brain injury. Behav Brain Res. 2007;181:200–209.

