## Supplement for "Effects of a single-dose methylphenidate challenge on resting-state functional connectivity in stimulant-treatment naive children and adults with ADHD"

### Supplementary Materials

#### Participants

Boys aged 10-12 years and men aged 23-40 years were included. Inclusion criteria were meeting criteria for a diagnosis of and requiring treatment with medication for ADHD (Inattentive, Hyperactive/Impulsive or Combined subtype). The diagnosis was determined by an experienced clinician based on the Diagnostic and Statistical Manual of Mental Disorders (DSM-IV; (American Psychiatric Association 1994)), which was confirmed with a (semi-)structured interview [1] in children; Diagnostic Interview for Adult ADHD (DIVA [2]). The DSM-IV requirement of at least six inattention or hyperactivity/impulsivity symptoms was applied to both children and adults. Participants were not eligible when they had received clinical treatment influencing the DA system (for adults before age 23), such as stimulants, neuroleptics, antipsychotics, D2/D3 agonists, or when they had a current or previous dependency on drugs that influence the DA system (for adults before age 23). Other exclusion criteria were an estimated IQ < 80 (Block Design and Vocabulary subtests of the WISC-III-R [3], Dutch Adult Reading Test [4], and/or a history of significant medical or neurological trauma or illness.

#### MRI acquisition

The MRI study was performed on a 3.0 T Philips scanner (Philips Healthcare, Best, The Netherlands) using an 8-channel receive-only head coil. Eight children and 1 adult were scanned on a 3T Philips scanner at a different center than the rest of the participants, also using an 8-channel head coil. A high-resolution 3D T1-weighted anatomical scan was acquired for registration purposes, and fMRI data were obtained using a single-shot echo-planar imaging sequence Parameters were: TR/TE=2300/30 ms, resolution=2.3×2.3×3 mm, 39 sequential slices, FOV=220x220x117 mm, GE-EPI read-out, no gap, 80° flip angle, 130 dynamics were used.

#### MRI preprocessing

Preprocessing was performed using FMRIPREP v1.2.3 [[5], RRID: SCR_016216]. Each T1w scan was bias-corrected, skull-stripped, and subsequently normalized to MNI space using non-linear registration. Functional data preprocessing included motion correction using FLIRT and distortion correction using an implementation of the TOPUP technique using 3dQwarp. This was followed by co-registration to the corresponding T1w using boundary-based registration with 9 degrees of freedom.

Motion correcting transformations, field distortion correcting warp, BOLD-to-T1w transformation, and T1w-to-template (MNI) warp were concatenated and applied in a single step using antsApplyTransforms (ANTs v2.1.0) with Lanczos interpolation. Independent component analysis (ICA) based on Automatic Removal Of Motion Artifacts (AROMA) was used to generate data that was non-aggressively denoised [6]. Two brain volumes were removed from the start of each scan to ensure that the steady-state equilibrium was attained.

Temporal signal-to-noise (tSNR) maps were calculated per participant to calculate the lowest-quartile mask per age group per session. Parcels overlapping with the lowest-quartile of the tSNR maps with more than 70% of voxels for more than 10% of the participants were excluded from the connectivity matrices [7].

Firstly, connectivity strength was measured, a measure of the temporal correlations in the blood oxygenation level-dependent (BOLD) signals between brain regions at rest. Secondly, two centrality measures were assessed, which reflect the influence a brain region has on the whole-brain network. We measured eigenvector centrality (EC), a measure of the extent to which a given brain region is connected to other highly connected regions, and betweenness centrality (BC), indicating the number of shortest connections that pass through it.

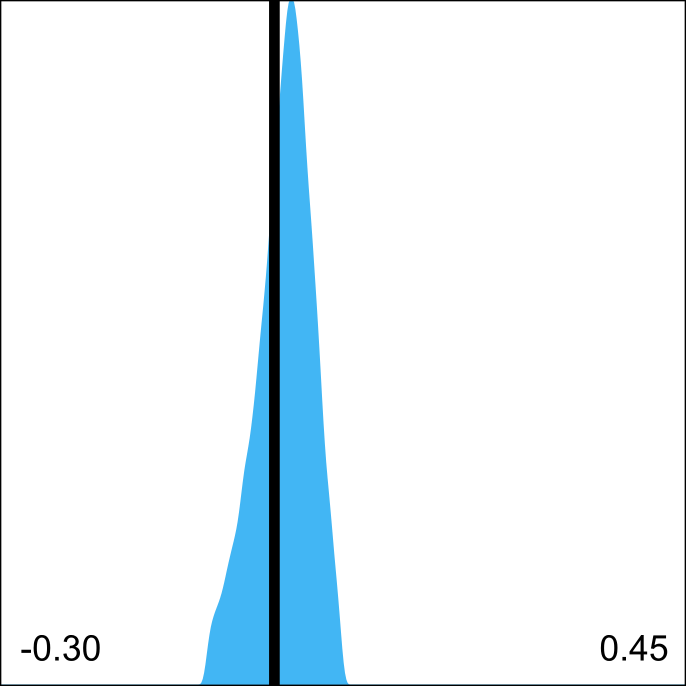

##### Supplementary Figure 1.| Residual QC-FC correlations after denoising. The distribution of all edgewise QC-FC correlations after denoising shows a narrow distribution and a distribution center close to 0 [8].

##### Supplementary Table 1.| Adjacency matrices - quality control.

| variable | Children | Adults |
| --- | --- | --- |
| # negative correlations | 7766.26 ± 4327.95 | 6638.26 ± 4386.57 |
| % negative correlations | 0.13 ± 0.07 | 0.11 ± 0.07 |
| Average Pearson's R | 0.31 ± 0.08 | 0.33 ± 0.10 |

##### Supplementary Table 2.| BNA parcellation numbers per region of interest (ROI).

| **Image** | **ROI** | **BNA atlas parcellation numbers** |
| --- | --- | --- |
| 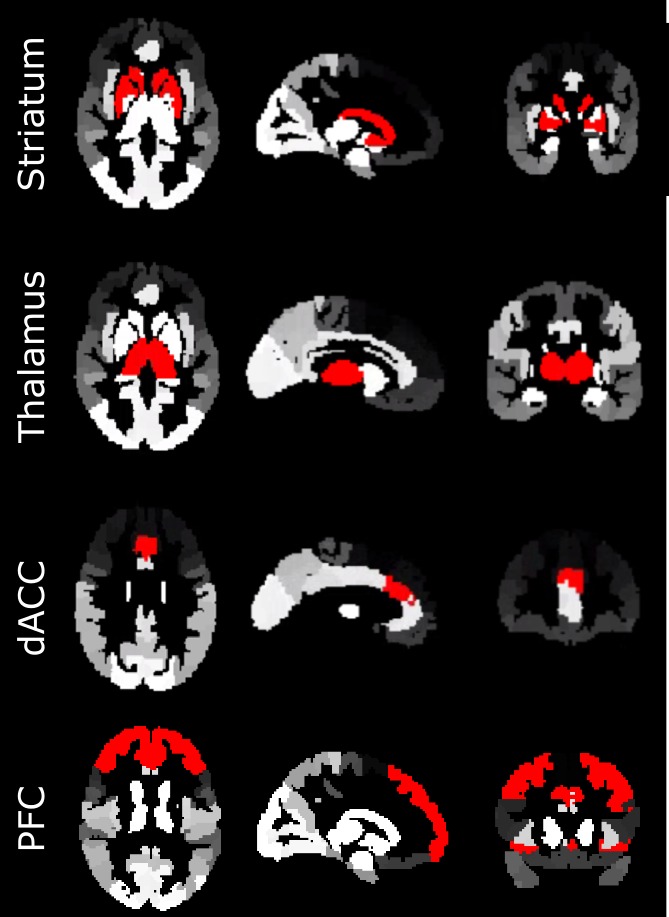 | **Striatum** | **219, 220, 225-230** |
| 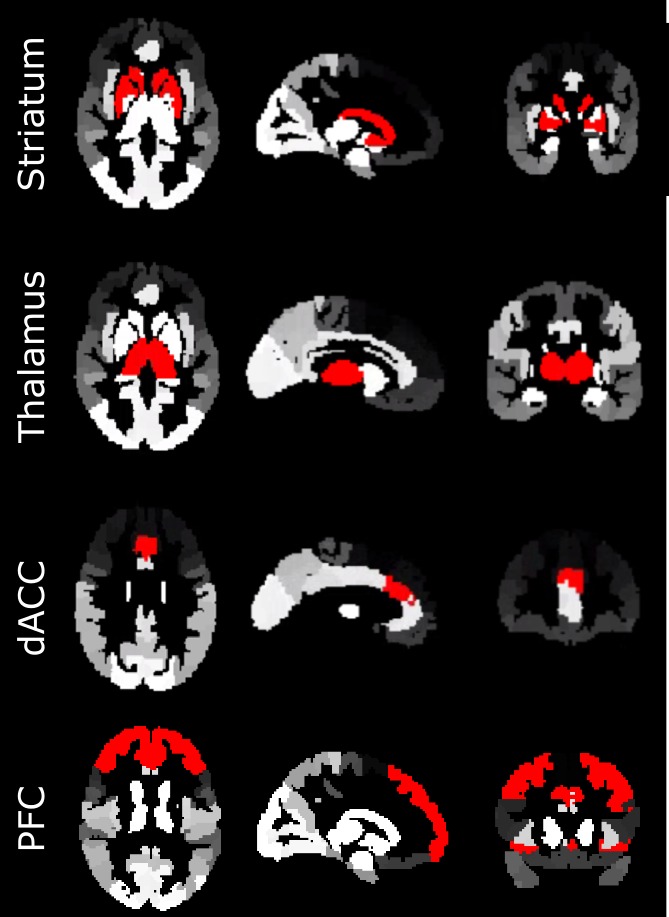 | **Thalamus** | **231-246** |
| 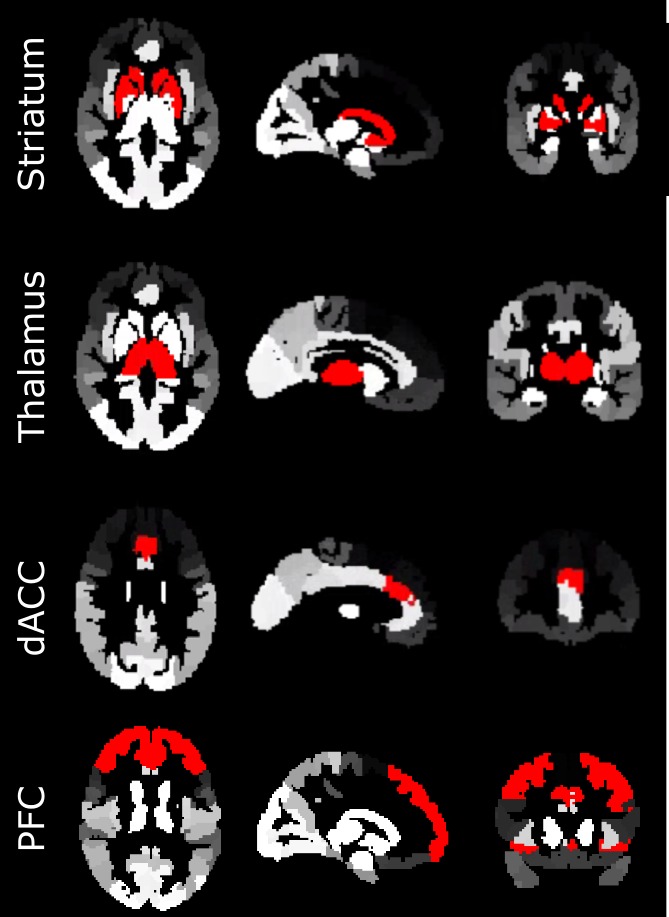 | **dACC** | **179, 180** |
| 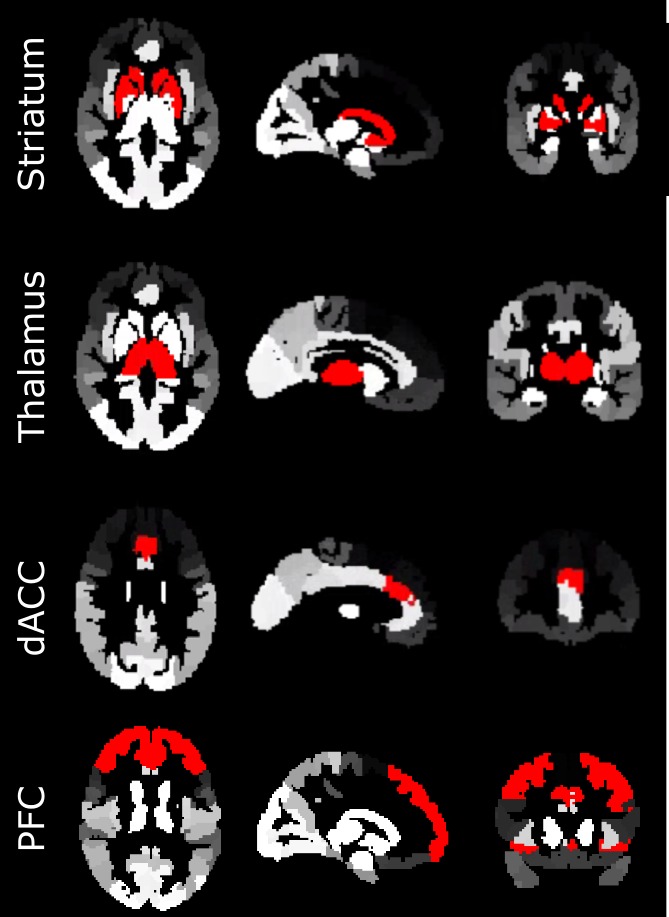 | **PFC** | **3-6, 11-28, 31, 32, 35, 36, 41, 42, 47, 48, 51, 52, 179, 180, 187, 188** |

### Supplementary Results

#### Participants

Included adult ADHD participants were significantly older than the TD adults (mean age ADHD= 28.5±4.6; mean age control 25.1±1.9; t(39)=3.90, p<0.01; but 80% of the adults with ADHD were under 30). TD controls did not differ from the ADHD participants in IQ (p>0.05). In addition, children and adults with ADHD differed from TD controls in ADHD symptom severity, as well as in anxiety and depressive symptoms.

Pre-methylphenidate, FD differed significantly between children and adults (ADHD: t(45)=5.48, p<0.01; controls: t(13)=4.99, p<0.01).

Excluded children did not differ in age, IQ, ADHD symptom-severity, anxiety symptoms, or depressive symptoms from the included children (p>0.05). Children with ADHD had significantly higher FDs than controls at pre-methylphenidate, but FD significantly decreased post-methylphenidate (F(1,87)=18.84, p<0.01). Pre-methylphenidate, adult ADHD FD values did not differ from controls, but they decreased post-methylphenidate (F(1,96)=10.08, p<0.01)(Supplementary Figure 2). Post-methylphenidate, FD of ADHD participants did not differ from controls. Importantly, except in two cases, (CS and BC in the PFC) none of the connectivity measures were correlated with motion (Supplementary Table 1).

##### Supplementary Table 3.| Correlation results of all connectivity measures with Framewise Displacement.

| variable | **Children**  BL | MPH | **Adults**  BL | MPH |
| --- | --- | --- | --- | --- |
| **STRIATUM** |  |  |  |  |
| strength | r=-0.07,p=0.60 | r=-0.24, p=0.12 | r=-0.12, p=0.38 | r=-0.20, p=0.18 |
| Eigenvector centrality | r=-0.01, p=0.94 | r=0.2, p=0.19 | r=-0.12, p=0.37 | r=-0.03, p=0.85 |
| Betweenness Centrality | r=0.23, p=0.08 | r<0.01, 0.98 | r=0.02, p=0.91 | r=-0.18, p=0.24 |
| **THALAMUS** |  |  |  |  |
| strength | r=0.01, p=0.92 | r=0.01, p=0.96 | r=-0.06, p=0.67 | r=-0.25, p=0.10 |
| Eigenvector centrality | r=0.17, p=0.21 | r=0.10, p=0.53 | r=-0.03, p=0.84 | r=-0.14, p=0.36 |
| Betweenness Centrality | r=0.18, p=0.17 | r=0.24, p=0.12 | r=0.23, p=0.09 | r<-0.01, p>0.99 |
| **DACC** |  |  |  |  |
| strength | r=-0.11, p=0.43 | r=-0.17, p=0.27 | r=-0.06, p=0.67 | r=-0.26, p=0.08 |
| Eigenvector centrality | r=-0.01, p=0.93 | r=-0.06, p=0.7 | r=-0.07, p=0.59 | r=-0.10, p=0.51 |
| Betweenness Centrality | r=-0.19, p=0.14 | r=-0.3, p=0.05 | r=-0.10, p=0.47 | r=-0.05, p=0.74 |
| **PFC** |  |  |  |  |
| strength | r=-0.10, p=0.46 | r=0.17, p=0.26 | r=0.52, p<0.01 | r=0.3, p=0.04 |
| Eigenvector centrality | r=-0.03, p=0.80 | r=-0.01, p=0.95 | r=-0.02, p=0.86 | r=0.16, p=0.28 |
| Betweenness Centrality | r=-0.02, p=0.86 | r=-0.29, p=0.05 | r=-0.58, p<0.01 | r=-0.25, p=0.08 |

##### Supplementary Figure 2.| Mean Framewise Displacement at pre and post MPH challenge for participants with ADHD (green) and control participants (grey). Showing estimated means with confidence intervals.

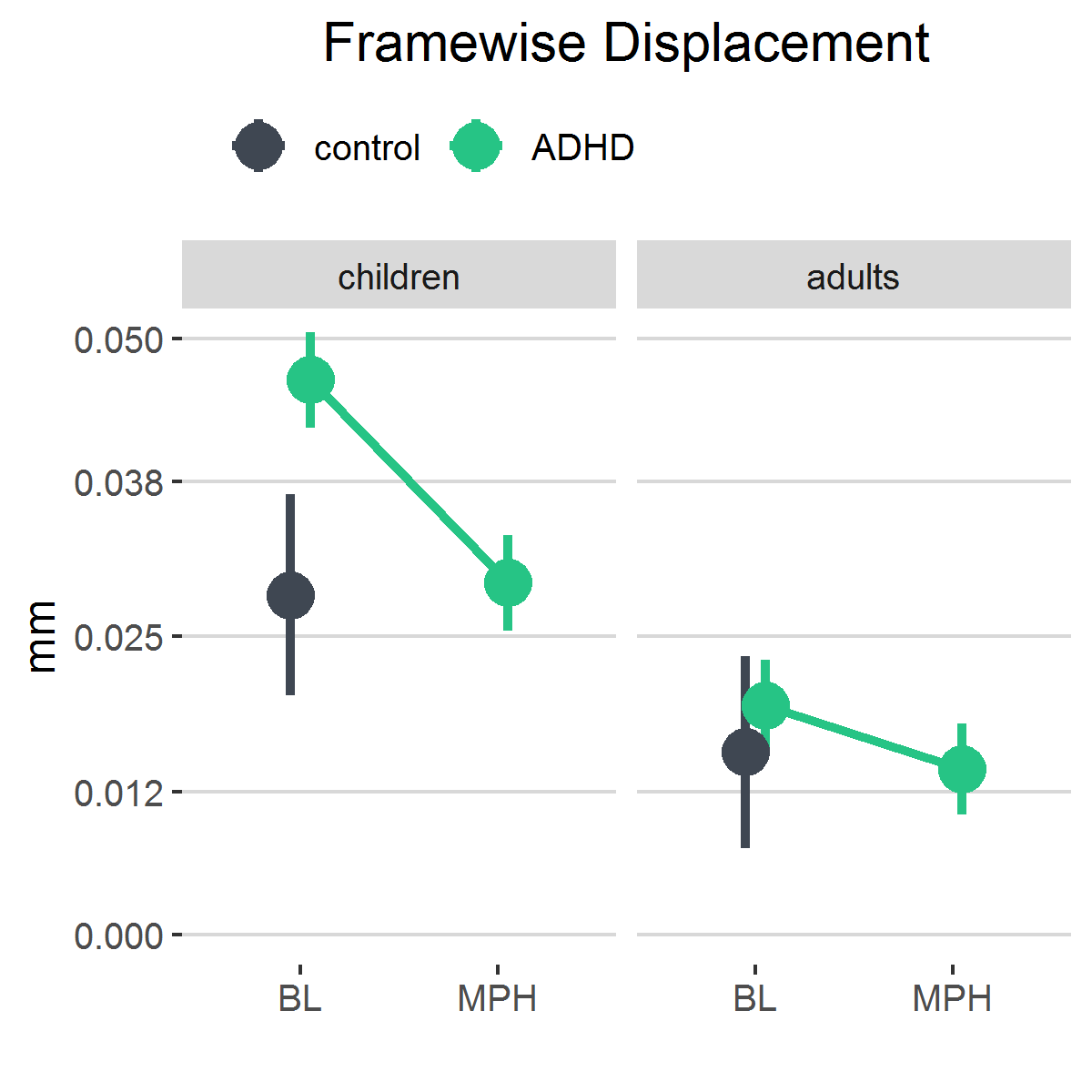

### References:

1.	Ferdinand R, van der Ende J. DISC-IV Diagnostic Interview Schedule for Children [Dutch translation NIMH-DISC-IV]. Rotterdam, the Netherlands: Afdeling Kinder- en Jeugdpsychiatrie, Sophia Kinderziekenhuis/Academisch Ziekenhuis Rotterdam; 1998.

2.	Kooij J. Adult ADHD: Diagnostic assessment and treatment. London: Springer-Verlag; 2012.

3.	Kort W, Compaan EL, Bleichrodt N, Resing WCM, Schittekatte M, Bosmans M. WISC-III NL. London Psychol Corp. 2002. 2002.

4.	Schmand B, Lindeboom J, van Harskamp F. Dutch Adult Reading Test. Lisse Swets En Zeitlinger. 1992. 1992.

5.	Esteban O, Markiewicz CJ, Blair RW, Moodie CA, Isik AI, Erramuzpe A, et al. fMRIPrep: a robust preprocessing pipeline for functional MRI. Nat Methods. 2019;16:111–116.

6.	Pruim RHR, Mennes M, van Rooij D, Llera A, Buitelaar JK, Beckmann CF. ICA-AROMA: A robust ICA-based strategy for removing motion artifacts from fMRI data. Neuroimage. 2015;112:267–277.

7.	Meijer KA, Eijlers AJC, Douw L, Uitdehaag BMJ, Barkhof F, Geurts JJG, et al. Increased connectivity of hub networks and cognitive impairment in multiple sclerosis. Neurology. 2017;88:2107–2114.

8.	Ciric R, Wolf DH, Power JD, Roalf DR, Baum GL, Ruparel K, et al. Benchmarking of participant-level confound regression strategies for the control of motion artifact in studies of functional connectivity. Neuroimage. 2017;154:174–187.
